# ANALYZING THE PATTERN OF USE OF ANTIBIOTICS IN A GHANAIAN DISTRICT HOSPITAL, ACROSS TWO DIFFERENT YEARS, USING THE WHO AWARE CLASSIFICATION

**DOI:** 10.1101/2023.10.27.23297681

**Authors:** Kwame Opoku-Agyeman, David Forson, Gerald Anim Acheampong, Daniel Addo, Thomas Afrifa Akwasi, Yaw Asiamah Asamoah, Ayim Britwum Prince, Adade-Boateng Kyei Baffour, Prince Boakye Frimpong

## Abstract

**Background:** There has been an increment in the resistance of microorganisms to various antibiotics used in regional hospitals in the Ashanti region of Ghana and the country. This has led to difficulty in selecting agents to treat patients without the associated increment in side effects, the astronomical increase in the cost of medical care and a longer duration of hospitalization. In this study, we analyze the pattern of antibiotic use in a district hospital using the WHO AWaRe recommendation.

**Objectives:** To ascertain the general pattern of antibiotic use with reference to the Access, Watch and Reserve Classes using the selected parameters (year of visit and patient demographics) and to confirm if the standard set by the WHO with respect to Access Class antibiotics is met.

**Method:** Files of patients, in 2021 and 2022, were systematically sampled from the database of SDA Hospital - Kwadaso (the hospital involved), and data concerning the visits for which antibiotics were administered were extracted to carry out the analysis required. The data extracted included the year of visit, demographics of the patients involved (patient status, age and sex) and the antibiotics that were administered.

**Results:** This study seeks to ascertain the distribution of antibiotics used across the two years in the hospital of interest using the 2021 WHO AWaRe (Access, Watch and Reserve Antibiotics) Classification. Ultimately, the value, in percentage, of the distribution of the antibiotics used in the Access class will be compared to the target of at least 60% of the total antibiotic consumption being Access group antibiotics as recommended by WHO.

## INTRODUCTION

Antibiotic resistance, an emerging global health issue, refers to the development of protective mechanisms to withstand the effects of antimicrobial agents, making antibiotics ineffective in hindering the growth of bacteria. (Beceiro et al., 2013; Nadeem et al., 2020). The emergence of Covid-19 has led to more frequent prescription and use of antibiotics to address secondary infections. This has prompted the World Health Organization (WHO) to express concerns that the efforts made in times past to fight antibiotic resistance may be rendered ineffective. (Getahun et al., 2020). Healthcare challenges associated with the pandemic such as increased hospital stays, overwhelmed staff, staff shortages and hindered infection control efforts, making it difficult to track resistant bacteria strains in healthcare facilities. (Afshinnekoo et al., 2021; Livermore, 2021).

The extensive use of antibiotics in not only healthcare but also agriculture has given rise to resistant microorganisms or strains in both humans and animals. (de Kraker et al., 2016; Pulingam et al. 2020; WHO, 2014). WHO estimates that by 2050, antimicrobial resistance could lead to 10 million deaths. Bacterial infections caused by resistant microorganisms usually lead to unfavourable clinical and economic outcomes such as prolonged illnesses, increased mortality rates, and soaring costs. These outcomes affect patient groups including the elderly, the immunocompromised and those in need of regular medical care. The transmission of antibiotic-resistant bacteria within healthcare facilities and the community is increasingly prevalent; raising a lot of concerns. (Farr et al., 2001; Suleyman et al., 2018).

Although antibiotics have demonstrated a lot of benefits in promoting human health by combating life-threatening bacterial infections, the development of antibiotic resistance hinders progress in safe food production, healthcare and life expectancy. This challenge along with the development of fewer new antibiotics over the years has worsened the issue. (Lewis, 2012; Singer et al., 2020). The improper use of new antibiotics can also reduce the efficacy of these antibiotics leading to short-lived responses. (Lee et al., 2013). The prolonged use of antibiotics has promoted the growth of resistant bacteria while inhibiting susceptible ones. Within the same class, continuous use of antibiotics can lead to resistance against multiple antibiotics. Previously, it was thought that maintaining resistance genes required additional energy and might not be stable under laboratory conditions, but recent findings contradict this notion (Davies and Davies, 2010; Melnyk et al., 2015).

There are several mechanisms bacteria employ in developing resistance to antibiotics. Bacteria employ various mechanisms to develop antibiotic resistance. This includes altering the antibiotic target site, inhibiting the antibiotic binding to the target site or modifying or destroying the antibiotic molecule. The development of resistance genes is through three main processes: transformation, transduction and bacterial conjugation. (Blair et al., 2015; Wright, 2010). Alternative antibiotics are being explored in the fight against antibiotic resistance. Some include antibiotic structure modifications, combinational antibiotic therapy, antibiotic-adjuvant combinations, aminoglycosides and their derivatives, and biopharmaceuticals. This review will expand on the economic ramifications of the intensifying antibiotic resistance crisis. It also highlights recent global illustrations of antibiotic resistance, and explains the mechanisms of resistance developed by bacteria and potential alternatives to antibiotics, underscoring the critical importance of antibiotic stewardship in safeguarding public health.

### Economic Point of View

From an economic point of view, antibiotic resistance presents a bleak outlook for the field of medicine. Global estimates predict losses ranging from $300 billion to $ 1 trillion. For OECD countries, cumulative economic output losses have been estimated to potentially reach $20-35 trillion. (Burki, 2018; O’neill, 2014). The economic burden in the United States stands at approximately $20 billion in healthcare costs and $35 billion in productivity losses (Ventola, 2015). The high rate of incidences of infectious diseases and the dependence on labour income have caused a disparate financial strain on the healthcare budgets of low-income countries. (Bank, 2017). Inadequate enforcement of regulations, the lack of education regarding responsible antibiotic use, and limited access to treatment exacerbate the issue in low- and middle-income nations (Bloom et al., 2017).

The economic ramifications of antimicrobial resistance include a potential 1% global GDP decrease, with developing countries facing a more significant 5-7% decline by 2050, worsening global inequality (Anderson et al., 2019; Bank, 2017). This will significantly affect low-income countries that depend on labor-based income. This will subsequently affect economic factors such as population size, human capital quality, and productivity levels. (Naylor et al., 2018). The number of exports is also affected especially in labor-intensive industries with declines anticipated by the year 2050. The global trade sector faces severe consequences in this regard. (Lekagul et al., 2019).

The development of antimicrobial resistance also increases mortality and morbidity among animals while hindering livestock production. (Hao et al., 2014). Antibiotic resistance in agriculture reduces the effectiveness of treatment. This drives up infection rates and prices of protein sources like meat, eggs and milk (Bank, 2017) and threatens the global supply of animal-based protein, crucial in meeting the demands of a growing global population (Van Boeckel et al., 2015). World Bank estimates suggest that low- and middle-income countries will face an 11% decline in livestock production by 2050, compounding economic challenges (Bank, 2017).

In 2014, antibiotic resistance claimed around 700,000 lives, with estimations indicating a potential global population reduction by 2050, with losses ranging from 11 million to 444 million people due to this global health crisis (O’neill, 2014). Disparities in antibiotic resistance patterns worldwide are linked to the misuse and overuse of antibiotics. Countries like India, Nigeria, Indonesia, and Russia, with high rates of infectious diseases like Malaria, HIV, or TB, are particularly vulnerable to worsening resistance (O’neill, 2014).

The commonly implicated antibiotic-resistant bacteria include P. aeruginosa, Enterococcus spp, A. baumannii, S. aureus, K. pneumoniae, and Enterobacter spp. Resistance extends to various antibiotics, as revealed in the WHO Global Antimicrobial Resistance and Use Surveillance System (GLASS) report 2021. E. coli and K. pneumoniae are prevalent, showing resistance to several antibiotics, including cefepime, ceftriaxone, ceftazidime, meropenem, and ciprofloxacin (WHO, 2021). The report throws light on E. coli strains resistant to 3rd generation cephalosporins, with noteworthy resistance in low-middle-income countries compared to high-income ones (WHO, 2021). Curbing antibiotic resistance is important in safeguarding public health and the global economy.

### Mechanism of antibiotic resistance

The issue of antibiotic resistance has been a global emergency for which solutions are being sought. That all antibiotics are susceptible to resistance while a few new ones are under development makes it a more pressing issue. Antibiotic resistance is a common phenomenon in developing countries and the emergence of different bacteria of varying resistance has been identified. (Basak et al., 2016; Magiorakos et al., 2012).

To appreciate how antibiotic resistance develops, it is essential to appreciate how antibiotics work. Antibiotics have active moieties which determine their efficacy. The deactivation of these active moieties is just one of the means through which they are rendered ineffective. On the other hand, microorganisms have developed structures and systems which grant them the capacity to overcome the activity of antibiotics.

Antibiotics work on microorganisms by targeting the following areas: nucleic acids, cell walls, protein synthesis and cell membranes (Kohanski et al., 2010).

### Biochemical aspects of resistance emphasize the various means through which microorganisms become resistant to antibiotics. These include

Efflux systems: Certain microorganisms, especially Pseudomonas aeruginosa, develop efflux systems or pumps that pump antibiotics out of their cells, reducing their effectiveness. The MexXY efflux system has been implicated in the development of resistance among Pseudomonas aeruginosa to aminoglycosides (Morita et al, 2012).

Microbial enzymes: The development of beta-lactamases by certain microorganisms gives them the ability to deactivate certain antibiotics, most of which belong to the beta-lactam class. The enzymes, called beta-lactamases cleave the beta-lactam ring found in these antibiotics, rendering them ineffective. Even relatively more reserved third-generation antibiotics such as cefotaxime are prone to this mechanism of resistance. (Chuma et al, 2013). Aminoglycoside-modifying enzymes are also produced by certain bacteria which limit their effectiveness. (Shi et al, 2013)

Decreased antibiotic penetration: Some bacteria also limit the penetration of antibiotics by decreasing the permeability of their cell membranes. For instance, Mycobacterium tuberculosis’s lipophilic outer cell wall hampers the entry of hydrophilic drugs.

Bypass of target sites and structures: Bacteria have the ability to generate substitute proteins to assume the function of the antibiotic’s intended target, thereby enabling them to withstand the drug’s impact. As an illustration, the mecA gene facilitates methicillin resistance in Staphylococcus aureus by encoding an innovative penicillin-binding protein (PBP) that supplants the drug’s original target (PBP-2A).

### The genetic features of certain microorganisms have been implicated in the development of antibiotic resistance. They include

Mutational resistance: The spontaneous mutations in the DNA of certain bacteria can confer antibiotic resistance. For example, mutations in the rpoB gene in Mycobacterium tuberculosis can confer resistance to rifampicin. (Pulingam et al, 2022) Gene transfer: Resistance genes can be transferred between bacteria through processes like conjugation, transformation, or transduction. This transfer of plasmids and transposons from one bacterium to the other can lead to the acquisition of resistance genes, contributing to the spread of antibiotic resistance. (Benett et al, 2009)

## ANTIBIOTIC STEWARDSHIP

Among the various classes of therapeutic drugs, antibiotics have proven to be a distinct drug class which possesses not only patient-centred outcomes but population or community-centred outcomes. The development of antibiotics has transformed medical practice significantly. This includes the ability to manage stubborn and dreadful infections (Doron S et al, 2011). The use of antibiotics has been on a continuous rise and this could be attributed to the increasing global population, increasing morbidity and rising number of new infections on a global scale. Globally, the use of antimicrobials has been said to increase by 36% from 2000 to 2010. This has led to a significant increase in global rates of infections characterized by resistant organisms and rising concerns about the narrowing of the antibiotic spectrum or coverage for these pathogens. This has potentially fatal public health consequences (Huttner et al.,2017; Schouten et al., 2017). Significant efforts to curb this worldwide issue by protecting the effectiveness of these antibiotics have been termed antibiotic stewardship.

Antibiotic stewardship refers to a coherent group of initiatives which optimize the sustainable use of antibiotics not only in human health but also in animal health or agriculture. There are core elements of antibiotic stewardship. These core elements are classified as four Ds: Drug, Dose, Duration, and De-escalation. These elements serve as principles that guide prescribers in choosing appropriate drug regimens which do not provoke resistance among various organisms for several antibiotics.

Antibiotic Stewardship has also been promoted through several interventional approaches such as broad or horizontal strategies and targeted or vertical level. Broad interventions are usually general strategies put in place to control the abuse of antibiotics at the prescription stage. Targeted or vertical strategies usually affect specific antibiotics or diseases. (Shrestha et al., 2023; Zahra et al., 2023) There can be mixed approaches where both horizontal and vertical strategies are put in place to reduce the inappropriate prescribing and use of antibiotics. At other health facilities such as community pharmacies and clinics, the use of empirical therapy consistently without investigations and the rising incidences of self-medication has led to a high number of poor outcomes stemming from resistant microbial species.

Antibiotic stewardship, in recent times, has been regarded to be an internationally recognized effort to improve various factors affecting the use of antibiotics including the prescribing and the judicious administration/use of antibiotics. It prioritizes key components including

- controlling the source of infection,
- enhancing infection prevention
- encouraging and supporting surveillance of antimicrobial resistance
- using the shortest duration of antibiotics based on evidence
- prescribing appropriate antibiotics with adequate dosages.
- reassessing treatment when culture results are available.

The goal of antibiotic stewardship is characterized by decreased rates of antibiotic resistance, improved clinical outcomes and cost-effective healthcare (Srinivasan A et al., 2017) and control Healthcare Professional Resources and Training programs need to be encouraged and organized to provide clinicians with continuous learning opportunities which will improve the frequency of use of antibiotics in the various groups: ACCESS, WATCH AND RESERVE.

In Ghana and other parts of Sub-Saharan Africa, the importance of antibiotic stewardship cannot be underestimated. The emergence of infections in these developing countries can be attributed to multifactorial causes including access to clean and portable water (especially in rural areas), crowded communities and slums, and poor sanitation in most communities, among others. Thus, the increasing incidences of infections including nosocomial infections implies that various antibiotics with diverse spectrum or coverage need to be prescribed and administered to manage these conditions. This has widely contributed to the development of resistance observed with several antibiotics including chloramphenicol. Antibiotic resistance has been regarded by many sections as a global epidemic with estimates of about 700,000 per year worldwide deaths attributable to antibiotic resistance. Projections of a million worldwide deaths by the year 2050 have also been made by statisticians based on the current trend and numbers observed. (Patel D et al., 2008)

In Ghana, the implementation of the “National Action Plan On Antimicrobial Resistance “has led to improved systems for data collection, analysis and interpretation of antimicrobial resistance. Various projects have been started to evaluate antibiotic resistance including studies on antibiotic resistance patterns in the elderly. There has also been the establishment of facilities including medical laboratories following the conclusion of these studies to facilitate antimicrobial resistance surveillance. Educational efforts have not been ignored in the attempt to create awareness of the long and short-term effects of antibiotic abuse and the factors that promote antibiotic resistance.

Workshops on antibiotic use have also been encouraged and patronized in most regional facilities in the country. These workshops have proven to be a good platform to advise prescribers on the need to avoid sticking with a select group of antibiotics, cephalosporin for example, to reduce the development of resistance in hospital and community settings.

The role of the media or press in promoting antibiotic stewardship cannot be underestimated. Media houses can make use of advertisements and programs to educate people on not only the impact of antibiotic resistance but also the potential side effects of and supply of restricted medicines. This requires that a restricted medicine including these medications with repeated exposure or intake.

Antibiotics are not supplied unless under a valid prescription. This has proven to be a good The Health Professions Regulatory Bodies Act 857 Section 4 places emphasis on the sale initiative to curb the abuse of antibiotics for conditions not indicated for and without confirmation of diagnosis from laboratory investigations. This has the potential to minimize the incidences of treatment failure in patients with confirmed hospital or community infections. This also possesses economic ramifications on the part of the system-level provider and beneficiary including reduced costs of healthcare.

Antibiotic stewardship requires a unified effort from all participants or stakeholders in the healthcare process. System-level providers, in particular, can influence the supply of medications which have been routinely and repeatedly used in a hospital facility.

Prescribers can also control the rate of antibiotic resistance and ultimately promote antibiotic stewardship. Patients or clients also play a role in antibiotic stewardship as they complete antibiotic treatment courses and avoid self-medication. Pharmacists, in particular, as stakeholders can make a huge difference in curbing the incidences of self-medication.

This is due to the high patronage of drugs from community pharmacies without any clear indication. In the Ghanaian setting, the use of chloramphenicol for cough in babies and toddlers is very common. Clinically, this can lead to a fatal syndrome or side effect known as the grey baby syndrome Pharmacists can intervene by questioning the direct purchase of antibiotics at the community pharmacy level before dispensing antibiotics. They can also educate clients or client relatives on the potential effects of drug use, particularly antibiotics without any clear indications for them. Pharmacists and prescribers can also reduce repeated exposure to antibiotics by reviewing the duration of therapy of clients on antibiotics. (Garau J et al., 2018)

Longer duration of treatment with antibiotics usually leads to higher eradication rates. For example, 14-day regimens of antibiotics in H. Pylori-induced peptic ulcer disease have proven to have better eradication rates than 10-day regiments. However, a longer duration of treatment does not always correspond with better clinical outcomes. Occasionally, tolerance and resistance are discovered with time due to repeated exposure. Also, in the case of antibiotic use in H. Pylori-induced peptic ulcer, clarithromycin resistance has been observed and noted globally which has led to the discovery of alternative regimens including the tetracycline therapy. (Braxton CC et al.,2010)

The issue of incomplete therapies is a major contributor towards the increasing rate of antibiotic resistance. In medication counselling sessions, prescribers need to educate patients or clients on the resistant species which can emerge from multiple incomplete drug therapies. Patients or clients are encouraged to complete antimicrobial drug courses or regimens to not only achieve positive clinical outcomes but also reduce the emergence of resistant species. (Lim CJ et al.,2014)

Alternative regimens can become a key cornerstone in the fight to protect the potency of antibiotics. Prescribers have an enormous role in avoiding skewed prescribing-sticking to a single antibiotic regimen for a particular type of infection. The use of alternative regimens reduces the exposure of an antibiotic to a particular microbial species. This makes it challenging for microbes to develop host responses to a single antibiotic and enables better clinical outcomes while reducing the need for repeated therapy. Thus, this emphasizes the need for educating prescribers and other health professionals on alternative regimens for diseases or infections. System-level interventions in this area can involve varying the supply of medications from regional stores to hospitals or clinics to broaden the scope of antibiotics prescribers can issue for therapy. This ensures that prescribers have a wide variety of medications to choose from.

The use of alternatives to conventional antibiotics should also be encouraged. Research promoting the discovery and exploring the specificity of antibodies as a solution to most infections should be supported and encouraged. Since microorganisms are unable to develop protective mechanisms against antibodies, this presents a therapeutic area with lots of potential for development. Also, the use of antimicrobial peptides like those found in casein is a potential replacement for conventional antibiotics. The low efficacy observed from natural antimicrobial peptides makes this a difficult approach for the drug. However, some scientists have suggested the potential use of synthetic peptides instead.

The use of bacteriophages in some parts of the globe with heightened success presents a possible therapeutic option for microbial infections. Bacteriophages are viruses that infect bacteria resulting in bacterial cell death. The main challenge with bacteriophages during therapy is the preparation of cultures for clinical use. Also, the transmission of genetic material implies that there is still some level of risk of bacteria resistance. Despite these challenges, bacteriophages have been proven to be more effective with wider spectrums than conventional antibiotics. The use of antibiotic oligonucleotides to prevent the expression of resistant genes and re-sensitizing resistant bacteria has also been suggested and is still being explored. (Trubiano et al., 2013)

Among these therapeutic approaches, bacteriophages and antibodies have been integrated into the healthcare process while the feasibility of the other options is still being explored. In cases where multi-drug resistant species these alternative options can be used either alone or in combination with conventional antimicrobials.

## METHOD

### Data Collection

From the Hospital Administration and Management Systems (HAMS) application, some patients’ files in 2021 and 2022 were systematically sampled, after which a Google form was created to extract the information needed. Within the form were the year of visit, the patient status (inpatient or outpatient), the sex (male or female), the age (<18 years, 18-64 years or >64years) and a checklist of antibiotics for which the administered antibiotics were selected or typed in. The extracted data was loaded into Microsoft Excel for analysis accordingly.

### Inclusion and exclusion criteria

- Patients’ folders from all wards and of all ages in 2021 and 2022 were used.
- Visits for which antibiotics were used were included.
- Visits or hospitalizations for which no antibiotics were used were excluded.

### Sample size

Out of a sample of approximately 110,000,400 samples would have been enough for statistical inferences; however, 783 files were selected to improve the power of this study.

### Sampling

A systematic sampling technique was employed in selecting patient folders from the HAMS for the studies: the 140^th^ file upon each count was selected in the entire list of files for both years, for the extraction of the required data, until the 783 responses were generated.

### Study Design

This is a retrospective descriptive study to elucidate the pattern of use of antibiotics using the AWaRe classification.

### Study Site

This study was carried out at SDA Hospital, Kwadaso – Ghana.

### Statistical Analysis

Antibiotics used in 2021 and 2022 were classified as Access-, Watch- or Reserve class antibiotics using the 2021 AWaRe Classification.

- The overall use of each class was expressed as a percentage of the total antibiotics for each stated year and plotted as bar graphs.
- Using patient demographics extracted, the relative distributions of antibiotics (in percentages) using the AWaRe Classification were plotted for each demographic.
- The specific antibiotics used in the hospital were classified and their extent of use, expressed in percentages of the grand total for each year, was also plotted.
- The top five indications and the proportions of their respective AWaRe antibiotics prescribed for them, in percentages, were represented on a bar chart.

### Ethics

The data collection process was done in line with the rules stipulated by SDA Hospital. Considering the retrospective nature of this study, each patient’s consent was practically impossible to obtain hence the informed consent of the patients involved was waived by the institution using its policies. However, confidentiality was strictly maintained and patients were kept anonymous throughout this study as evidenced by the demographics extracted, which were all non-specific.

## RESULTS

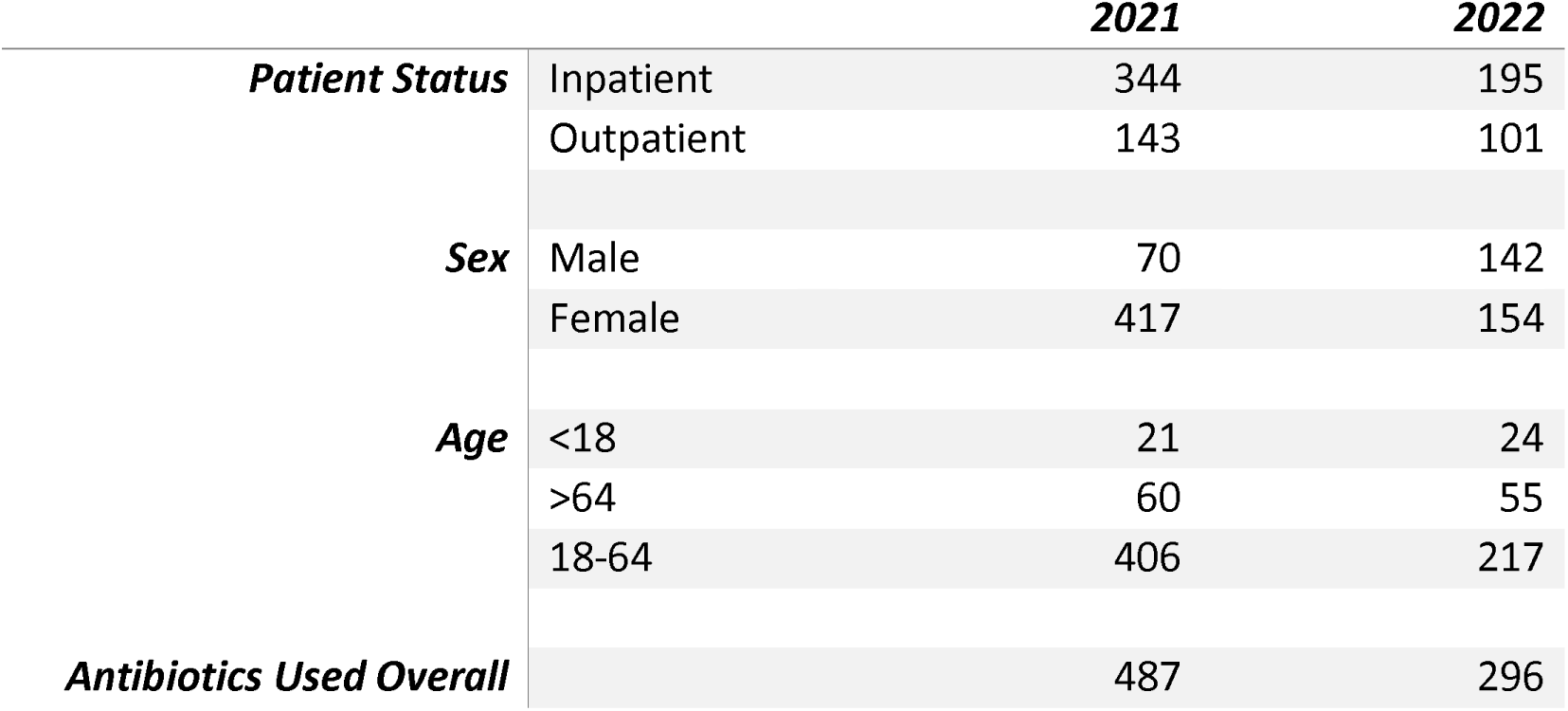

### PATIENTS’ DEMOGRAPHICS

#### Patient Status

For 2021, 344 inpatients and 143 outpatients were involved. For 2022, 195 inpatients and 101 outpatients were involved.

#### Sex

For 2021, 417 of the patients involved in this study were females with 70 being males. For 2022,154 of the patients involved in this study were females with 142 being males.

#### Age

For 2021, 21 of the patients involved in this study were < 18 years of age, 60 were >64 with 406 falling within the range of 18-64. For 2022, 24 of the patients involved in this study were < 18 years of age, 55 were >64 with 217 falling within the range of 18-64.

#### Overall Use of Antibiotics

The total number of antibiotics prescribed in 2021 per the sample used here was 487 (Access – 228, Unclassified – 20 and Watch – 239). For, 2022 per the sample used here the total number was 296 (Access – 151, Unclassified - 3 and Watch – 142).

### DISTRIBUTION OF ANTIBIOTICS USING THEIR RESPECTIVE TRADITIONAL CLASSES

As shown in Figure 0, in 2021 and 2022 respectively, tetracyclines made up 0.6% and 1% of the total antibiotics used. Sulfonamides made up a corresponding 1.0% and 0.7% whereas penicillins made up 18.9% and 16.9% of antibiotics used respectively. Nitroimidazoles covered percentages of 20.3 and 23.6 whereas macrolides, 7.0 and 5.7 in 2021 and 2022 respectively. 8.1 and 4.9 were the percentages obtained for the use of lincosamides in 2021 and 2022 respectively. Fluoroquinolones made up 11.5% and 16.9% whereas cephalosporins made up 34% and 26.4% of antibiotics used in 2021 and 2022 respectively. Lastly, aminoglycosides covered percentages of 1.8 and 0.7 for antibiotics used in 2021 and 2022 respectively.

### DISTRIBUTION OF ANTIBIOTICS USING AWaRe CLASSIFICATION

For patient status, as shown in Figure 1, 48% of the antibiotics used fell within the Access Class whereas 48% fell within the Watch Class for inpatients. The unclassified class accounts for the remaining percentage. For outpatients,46% of the antibiotics used fell within the Access Class, 49% within the Watch Class with the remaining 5% falling in an Unclassified Group using the 2021 AWaRe Classification. For 2022, as shown in Figure 2, 56% of the antibiotics used fell within the Access Class whereas 44% fell within the Watch Class for inpatients. For outpatients, 42% of the antibiotics used fell within the Access Class, 57% fell within the Watch Class and 1% of the antibiotics used were found to be unclassified.

**FIGURE 1.**
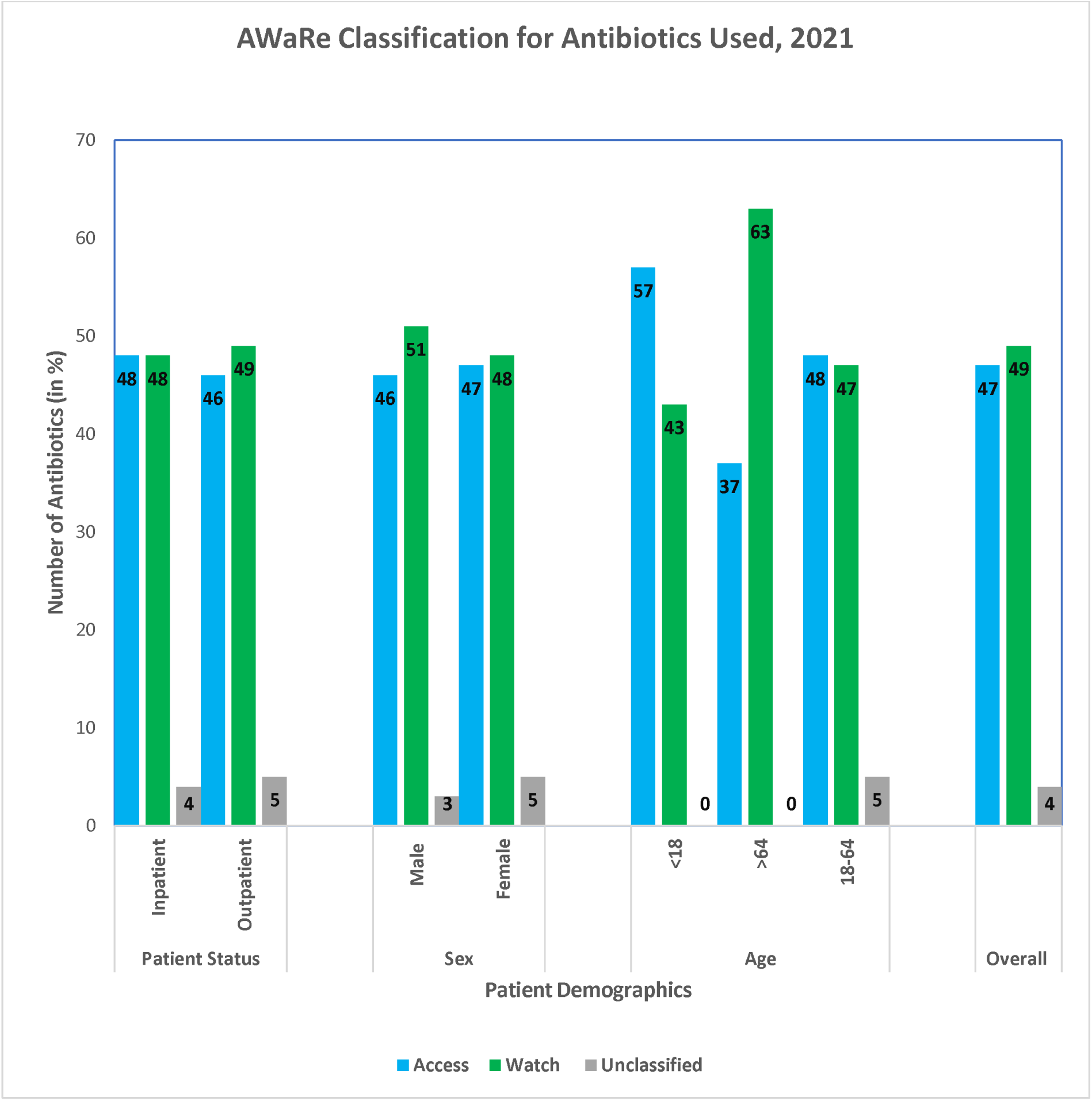
REVIEW OF ANTIBIOTIC USE, 2021.

**FIGURE 2.**
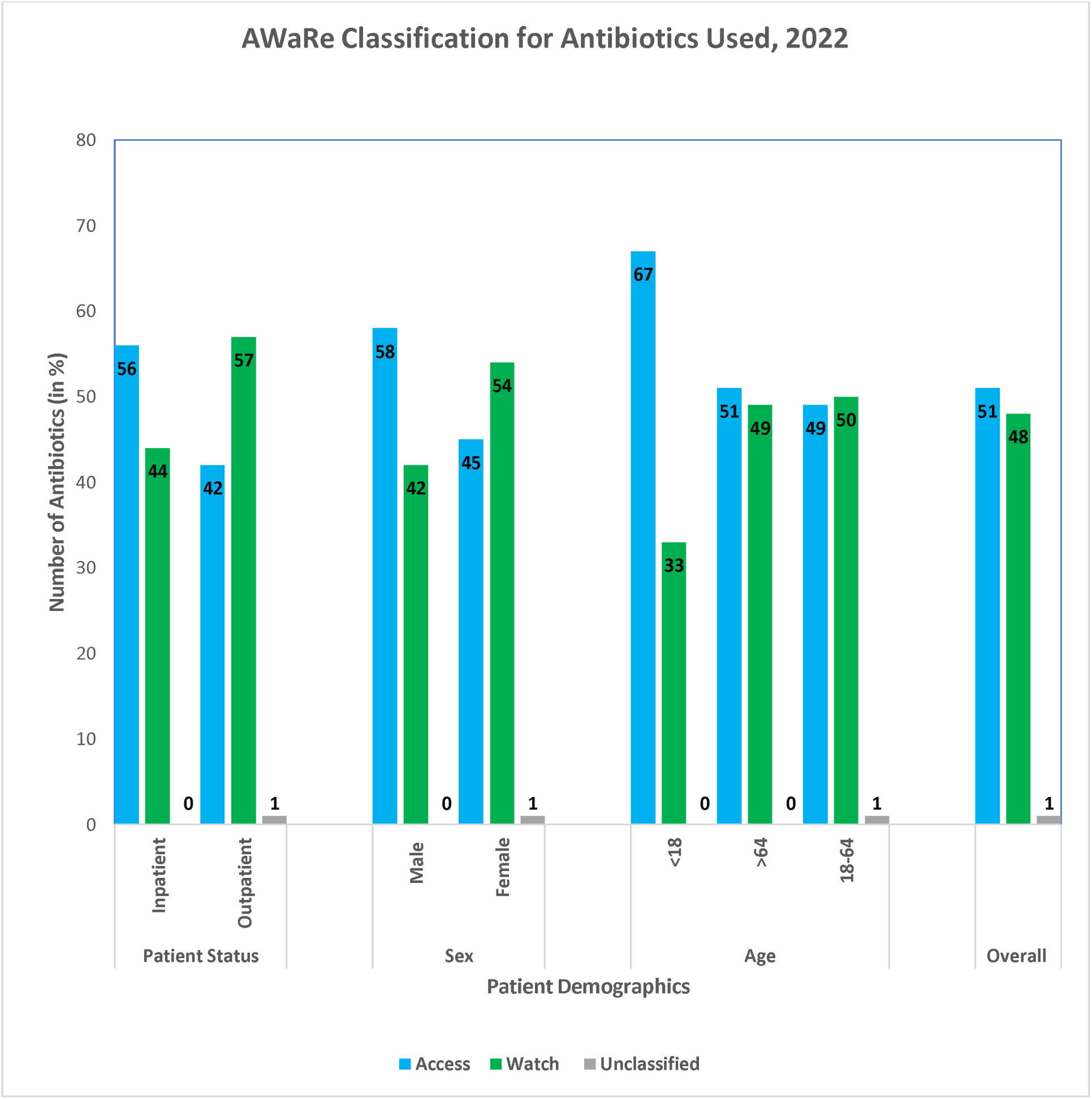
REVIEW OF ANTIBIOTIC USE, 2022.

Using the sex demographic, as shown in Figure 1, for females, 47% of the antibiotics used were Access class antibiotics, 48% were Watch Class and 5% made up the Unspecified Class. 46% of the antibiotics were Access Class, 51% were Watch Class Antibiotics for the males whereas 3% were of the Unclassified class. For 2022, as shown in Figure 2, for females, 45% of the antibiotics used were Access class-, 54% were Watch Class- and 1% were Unclassified antibiotics. 58% of the antibiotics were Access Class and 42% were Watch Class Antibiotics for the males.

For the age distribution, as shown in Figure 1, For patients <18, 57% of the antibiotics used were Access class with the remaining belonging to the Watch class. For patients >64, 37% of the antibiotics used fell within the Access class, whereas the remaining fell within the Watch class. For patients 18-64 years of age, 48% of the antibiotics used were Access class antibiotics, 47% were Watch class antibiotics with the remaining 5% belonging to an unspecified group. For 2022, as shown in Figure 2, for patients <18, 67% of the antibiotics used were Access class with the remaining belonging to the Watch class. For patients >64, 51% of the antibiotics used fell within the Access class, whereas the remaining fell within the Watch class. For patients 18-64 years of age, 49% of the antibiotics used were Access class-, 50% were Watch class and 1% were Unclassified antibiotics.

The Access-, Unclassified- and Watch groups in 2021 had percentages of 47, 4 and 49 respectively as shown in Figure 1. No Reserve class antibiotic was recorded for the sample used in this study. For the antibiotics used in 2021 as shown in Figure 3, the most frequently used antibiotics were Metronidazole, Cefuroxime and Amoxicillin + Clavulanic Acid. Ciprofloxacin, Ceftriaxone, Azithromycin and Clindamycin came second in rank with respect to their frequency of use with the remaining antibiotics being barely used. The Access-, Unclassified- and Watch groups in 2022 had percentages of 51, 1 and 48 respectively as shown in Figure 2. None of the antibiotics used in 2022 fell within the Reserve Class antibiotics for the sample employed in this study. From Figure 3, the most frequently used antibiotic was Cefuroxime. Ciprofloxacin, Metronidazole, Ceftriaxone and Amoxicillin + Clavulanic Acid were the most employed antibiotics after Cefuroxime. Clindamycin and Azithromycin were moderately used whereas the remaining antibiotics were barely used.

**FIGURE 3.**
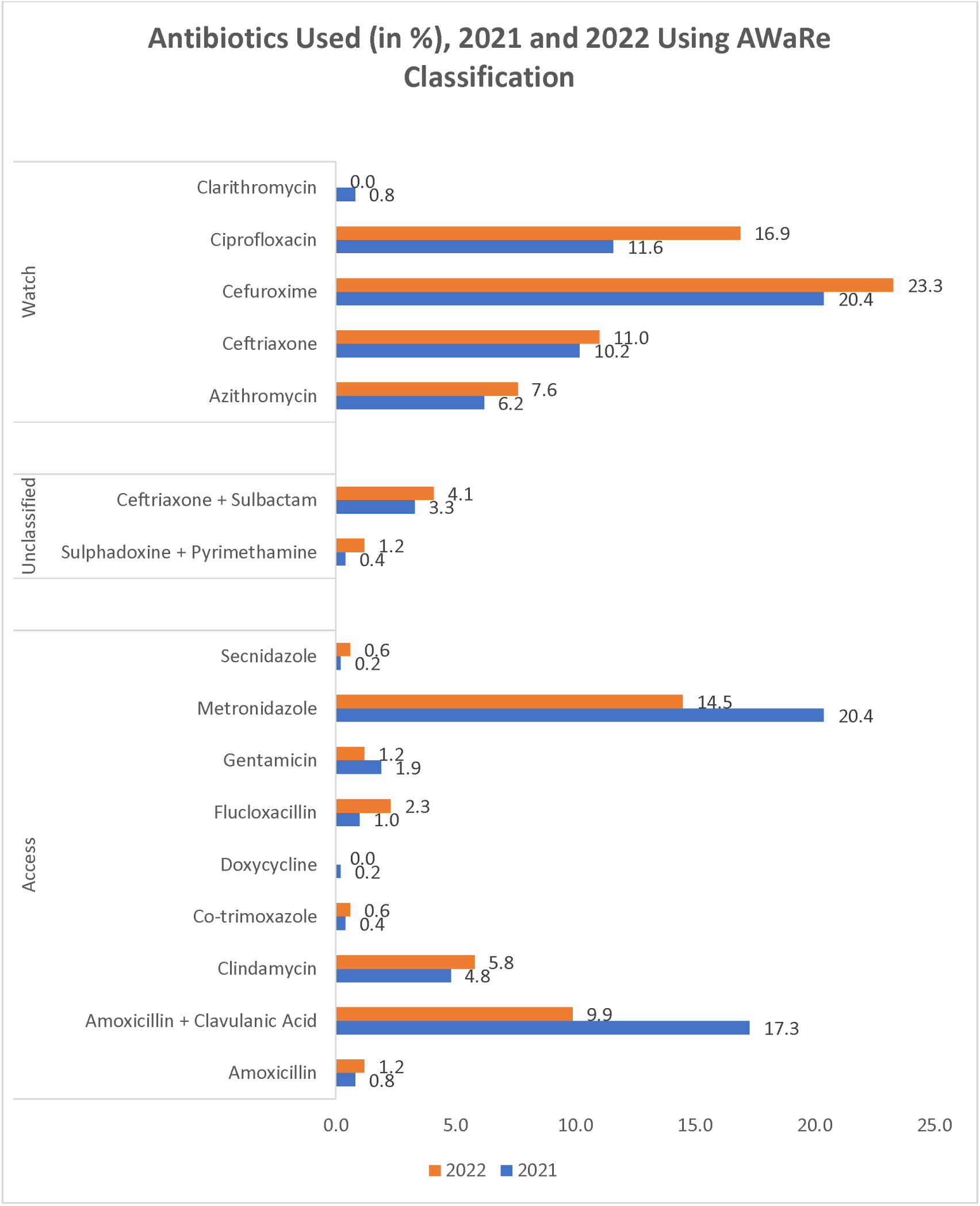
ANTIBIOTICS USED IN 2021 AND 2022.

**FIGURE A.**
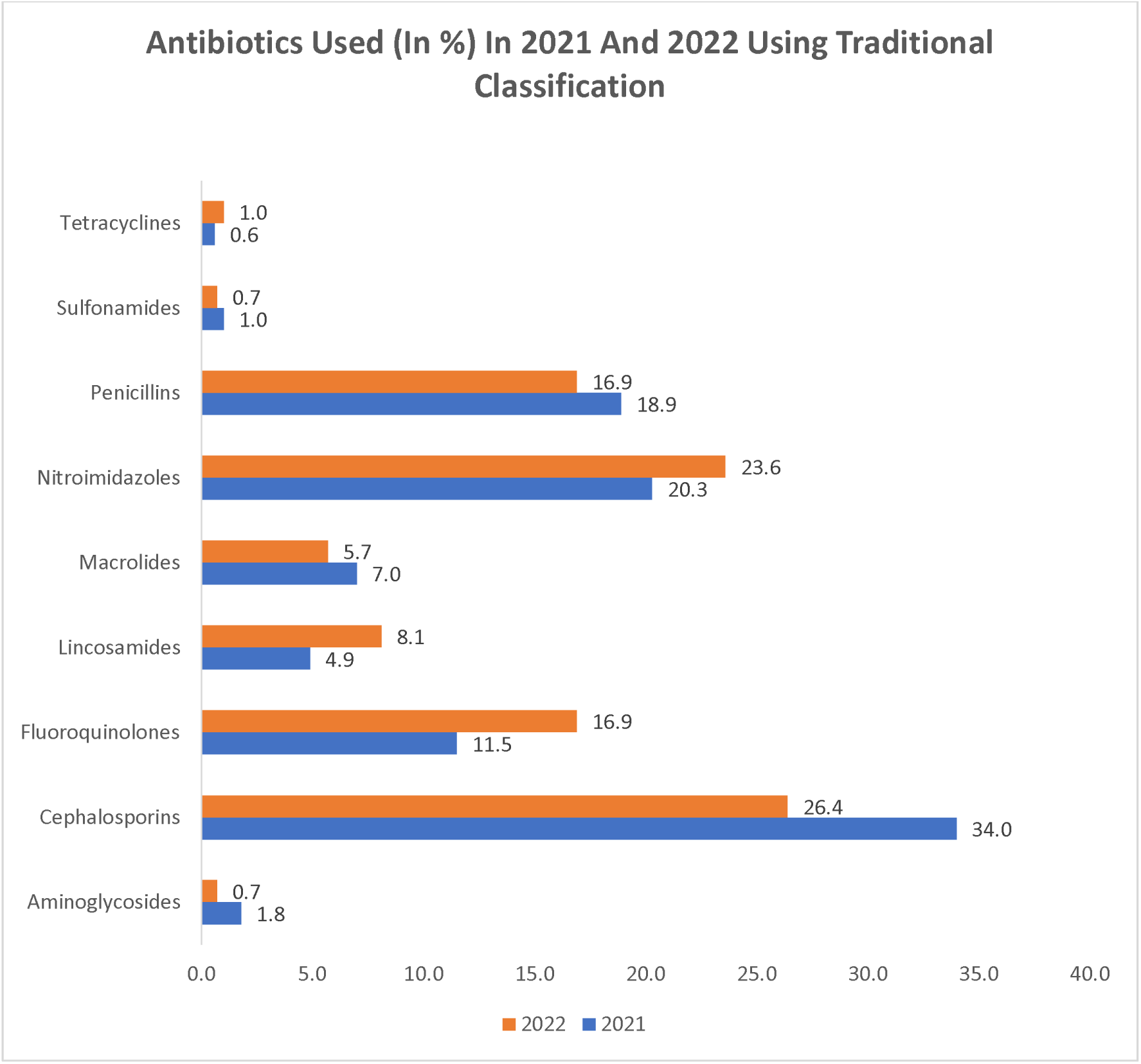
DISTRIBUTION OF ANTIBIOTICS USING THEIR RESPECTIVE TRADITIONAL CLASSES.

**FIG B.**
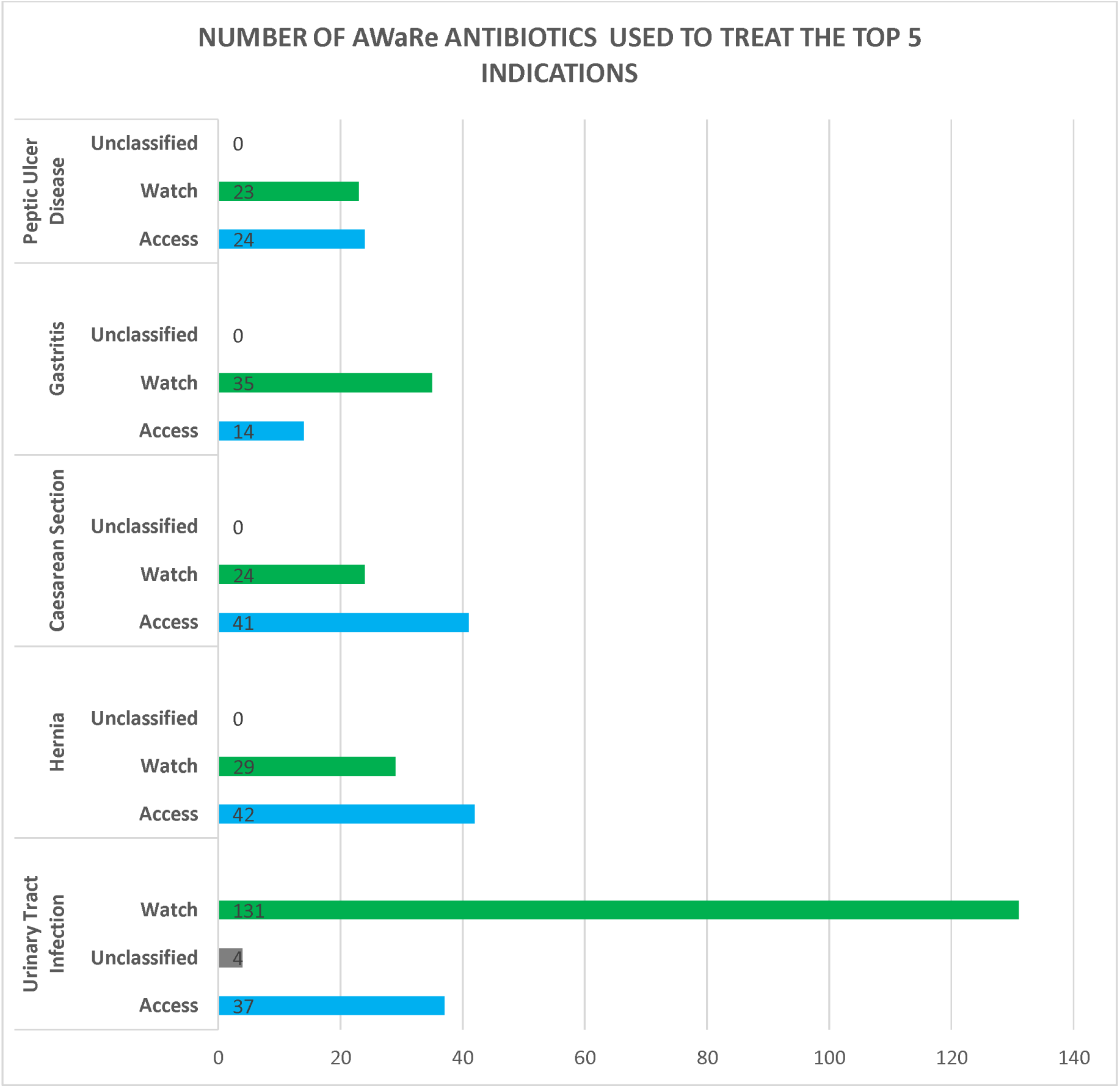
DISTRIBUTION OF AWARE ANTIBIOTICS AMONG THE TOP FIVE INDICATIONS IN THE HOSPITAL.

## DISCUSSION

The concerns about antimicrobial resistance (AMR) and its ubiquitous impact on several sectors, of which the medical sector is the most crucial, have necessitated the judicious and rational use of antibiotics. On the impact of AMR on health, it’s been emphasized that the cost of medical care, the length of hospitalizations and the toxicity of the more reserved groups would be a huge blow to the various efforts to improve the accessibility of quality yet affordable healthcare.

This led to the development of the 2021 WHO AWaRe classification to ensure that a standard is set to guide and monitor the rational use of antibiotics, which will in turn help mitigate antimicrobial resistance (AMR). Under the 2021 AWaRe classification, antibiotics are classified into three groups: Access, Watch and Reserve. The Access class of antibiotics is recommended as the first-line drug for the treatment of commonly encountered infections since they have a lower resistance potential relative to the other drugs. On the other hand, the Watch class of antibiotics is typically recommended as second-line agents, after the Access antibiotics have failed, but can be used as first-line agents based on the type of infections and the resistance profile of the facility involved. The Watch class have a higher resistance potential than the Access class. Finally, the Reserve class of antibiotics is used as the last resort for the treatment of infections and is employed when all other agents have failed. They are used typically in the management of multi-drug-resistant organisms.

The AWaRe classification defines targets for the rational use of antibiotics at the local, national and global to improve antimicrobial resistance. On this wise, a country-level target of at least 60% of antibiotics used is expected to fall within the Access Group. For this target to be achieved at the national level, it surely must be met at the institutional level. In this study, we analyzed the use of antibiotics in 2021 and 2022 by classifying the various antibiotics used in the samples obtained and quantifying them to ascertain if they met the institutional level of 60%, by extension of the national-level target. Of the 783 samples employed, the antibiotics used were classified under the Access, Watch and Reserve Groups. Amoxicillin, Amoxicillin Clavulanic Acid, Clindamycin, Co-trimoxazole, Doxycycline, Flucloxacillin, Gentamicin, Metronidazole and Secnidazole made the Access group up whereas Azithromycin, Ceftriaxone, Cefuroxime, Ciprofloxacin and Clarithromycin constituted the Watch group. Two agents were found not to exist within the three classes and were grouped as “Unclassified” and were made up of Sulphadoxine Pyrimethamine and Ceftriaxone Sulbactam.

The use of these antibiotics was assessed over three patient demographics: Patient Status (Inpatients and Outpatients), Sex (Male and Female) and Age (>18years, 18-64years and >64years) after which the total number of antibiotics for each class was expressed as a percentage of the grand total and compared to the target set by the WHO AWaRe classification. Since the WHO AWaRe Classification targets the Access class, our discussion will focus on and compare the Access values of each demographic in 2021 and 2022. In 2021, using patient status, both inpatients and outpatients had 48% and 46% respectively compared to the 56% and 42% in 2022 respectively. For sex, both males and females had percentages of 46 and 47 in 2021 respectively whereas 58 and 42 were the corresponding values obtained in 2022. On the premise of age, patients > 18 years, 18-64 years and > 64 years had percentages of 57,37 and 48 respectively while 67, 51 and 49 were the corresponding values in 2022. Overall, the total number of Access antibiotics used in 2021 and 2022, expressed as a percentage of the total number of all antibiotics used (Access, Watch and Unclassified) were 47 and 51 respectively. This brings to light that the institutional level of 60% for Access Class Antibiotics is not being met widely even though the target was achieved for patients < 18 years of age in 2022. As demonstrated in another research project in Ghana, a huge amount of antibiotics used falls within the Watch class (1). This is further seconded by another study that emphasizes the high rate of prescription of Watch class antibiotics in low- and middle-income countries, of which Ghana forms a part. (6) The high rate of prescription of Watch antibiotics could be attributed to the resistance patterns observed in the hospital, the knowledge of the prescribers and the availability and affordability of certain antibiotics. For instance, in patients less than 18 years, across both years, the Access group are the most prescribed and has been found to be effective in curing ailments in such population; however, in patients greater than 18 years, the Watch group tends to be relatively prescribed the more. This could be attributed to the concept of “Adaptive mutation”, where because of prior exposure to several antibiotics for patients in the latter group either as inpatients or outpatients, the pathogens may have been exposed to sub-inhibitory levels of the involved antibiotics - on the premise of medication non-compliance particularly for outpatients - which leads to antimicrobial resistance. Hence, the Watch class antibiotics are resorted to for most indications among such groups on a “tried-and-tested” basis.

Metronidazole was the most used antibiotic in the Access group followed by Co-amoxiclav whereas Cefuroxime was the most used antibiotic among the Watch group followed by Ciprofloxacin, as shown in another Ghanaian-based study (2). Another point to note is that the most used antibiotics were cephalosporins as shown in a similar study (4). We also moved on to take a look at the top five indications in the facility for which the various antibiotics were used the most. The indications, in order of ranking, were Urinary Tract Infection, postoperative management for Hernia and Caesarean Section, Gastritis and Peptic Ulcer Disease. For Urinary Tract Infections and Gastritis, more Watch antibiotics were used relative to the Access whereas the converse was true for the remaining three. The stark difference in the number of antibiotics between the Access and Watch groups in the treatment of Urinary Tract Infections raises a lot of concern and hints at the increased resistance of uropathogens cross-continentally in select populations to Access antibiotics as shown similarly in another research in Hungary. (3)

The difficulty here has to do with defining the appropriateness of the pattern of antimicrobial use in this setting since factors such as the resistance patterns in the hospital, and the cost and availability of the medication used all have a role to play in the overall outcome. To improve the usage of antibiotics within the Access class, we believe organizing awareness programs for healthcare professionals on the use of antibiotics for Access class-susceptible infections coupled with developing a system to monitor and report adherence to the set guidelines will be the first step in the right direction. This vision is shared by another study that assessed the impact of raising the awareness of healthcare professionals on the WHO Aware classification (5). Another sustainable method will be to monitor the resistance patterns within the facility, develop antibiograms and update the guidelines appropriately and timely in order to make them relevant and workable.

To our knowledge, this is the first study that analyzes the pattern of use of AWaRe antibiotics using multiple patient demographics, such as patient status, age and sex simultaneously across two different years. The strength of this study lies in the multiple determinations of the proportion of AWaRe antibiotics used for each patient demographic and the overall antibiotic usage. This reveals patterns at multiple levels and informs decisions to be made at each of these levels. On the flip side, our study fails to capture the duration and dose of antibiotic use, the wards at which they were prescribed and the clinical outcome. This study also fails to capture the precise reasons for the pattern of antibiotic usage for the various demographics, given that the reasons stated were based on a limited survey. Our study was done in one hospital which also limits its generalizability. On this note, we propose a future study to ascertain the reasons for the pattern of antibiotics usage using the AWaRe classification coupled with the study of the distribution of AWaRe antibiotics on a unit-by-unit (such as Surgical, Maternal, Internal Medicine etc.) basis to tailor the development of antibiotic protocols at each unit level. Finally, carrying out similar studies at various hospitals will provide a more robust and refined look at the pattern of antibiotic usage on a regional, national and even global level.

The AWaRe classification is a tool we believe could be used as a prototype to develop institution-specific antibiotic use guidelines. This would factor in concerns such as the resistance patterns, and the cost and availability of antibiotics to foster a desirable use of antibiotics at the local level and subsequently, at the national level.

## CONCLUSION

There was a low Access class antibiotics usage-47% and 51% in 2021 and 2022 respectively-compared to the 60% standard set by the WHO in the hospital. Developing a robust institution-specific system hinged on the WHO AWaRe Classification, which also factors in local driving forces, to guide and inform clinicians on the use of antibiotics will guide their rational use and decrease the incidence of antimicrobial resistance.

## Data Availability

All data produced in the present work are contained in the manuscript.

## Abbreviations

AMR: Antimicrobial Resistance
AWaRe: Access Watch and Reserve Classes of Antibiotics
WHO: World Health Organization

